# Experiences and Perceptions of Regular Floatation-REST Users: Results from an Online Survey

**DOI:** 10.1101/2025.04.14.25325793

**Authors:** Kai Hilpert, Irina Chis Ster

## Abstract

**Objective:** Since the 1980s, flotation Restricted Environmental Stimulation Therapy (REST) has gained international recognition, particularly with the advent of modern flotation tanks using Epsom salt for buoyancy. Concurrent clinical studies have demonstrated the efficacy of flotation REST in treating various conditions, including depression, anxiety, and pain. Despite its growing popularity, the motivations, experiences, and insights of regular flotation REST users remain underexplored. This study aims to investigate these aspects in detail.

**Design:** Between 6th January and 30th April 2023, a comprehensive online questionnaire was administered. The 23 questions covered various aspects of flotation practices, such as frequency, average duration, and perceived benefits, including stress reduction, sleep improvement, and pain management. Demographic data, including gender, age, and occupation, were also collected. Additionally, participants were invited to suggest potential areas for future research into flotation-REST therapy.

**Results:** In total 83 regular floating users provided an answer, stemming from the following countries, the UK (52), Canada (7), Australia (6), New Zealand (5), Ireland (4), Belgium, Serbia and Portugal (each 1) and 1 undeclared. The majority of participants were female (63.4%), with a significant number aged between 26 and 55 years. About 70% of participants reported they would prefer to float 1-7 times a week if not limited by time or money, though current usage was mostly 1-3 times a month, coinciding with typical membership plans at float centers. The survey revealed that about 65.1% of participants typically engage in flotation sessions lasting 30 to 60 minutes, while 28.9% opt for sessions between 61 to 90 minutes. In terms of experience with regular flotation, the largest group, accounting for 26.5% of respondents has been practicing for 3-5 years. The perceived benefits of floating were high levels of relaxation (91.6%), stress reduction (77.1%) and time without external stimulus (67.5%). Other significant benefits included mental clarity (56.6%), sleep improvement (55.4%), anxiety (47.0%), pain management (33.7%) and depression (21.7%). Furthermore benefits of personal growth were noted, for example a deeper connection to self (53.0%) and spiritual development (44.6%). The majority of participants (53.0%) experienced those benefits for 1 - 4 days, followed by 33.7% where the benefits lasted between 2 - 23 hours. The majority of participnats were very supportive of new research, for example, the majority of respondents (84.1%) recommended research on stress reduction in general. Other significant areas of interest include burn-out prevention, trauma/Post-traumatic stress disorder (PTSD),each receiving 73.2% of responses, and anxiety (70.7%). Reported side effects (25.5%) were all very minor.

**Conclusion:** Flotation therapy is shown to be highly valued by its regular users for its mental health benefits, notably in stress reduction and relaxation. The desire for more frequent sessions suggests that users perceive ongoing benefits that could be optimized through regular participation. The study underscores a significant interest in further research, particularly in enhancing the understanding of flotation therapy’s impacts on pain management, sleep quality, and mental health conditions like PTSD. These findings advocate for a broader public awareness and deeper scientific exploration to harness and expand the therapeutic potential of flotation therapy, making it a more accessible and recognized wellness tool.

## 1. Introduction

### 1.1 Floatation-REST history

The origins of Floatation REST can be traced back to the mid-20th century, with the pioneering work of John C. Lilly and Jay T. Shurley, who developed the first floatation tank in 1954 ^1–3^. Lilly’s initial experiments were aimed at studying the effects of sensory deprivation on the human brain and consciousness. His initial experiments involved vertical tanks filled with warm water and sodium chloride (NaCl) ^1^. These early tanks were basic, focusing on isolating the individual from external stimuli. The transition from sodium chloride to magnesium sulfate (Epsom salt) emerged as practitioners sought to enhance the floating experience. Epsom salt provided greater buoyancy, allowing for more effortless floating and reducing the risk of saltwater discomfort ^4^.

In the 1970s, Glenn Perry, in collaboration with his wife Lee Perry and Dr. Lilly, designed the first commercially available floatation tank. Their company, Samadhi Tank Company, was instrumental in making horizontal floatation tanks accessible to a broader audience ^4^. The evolution of floatation tanks has seen significant advancements in design and technology, with modern tanks featuring temperature control, light and sound customization, ergonomic features, advanced filtration systems, and improved aesthetics and accessibility.

Peter Suedfeld’s research in Restricted Environmental Stimulation Therapy (REST) played a crucial role in establishing floatation REST as a legitimate scientific and therapeutic practice ^5–10^. Additionally, researchers like Thomas H. Fine and Roderick Borrie in the 1980s conducted significant studies on the therapeutic and relaxation benefits of floatation REST, further legitimizing the practice^11–16^. Over the decades, the practice evolved from a scientific tool to a therapeutic technique, gaining popularity in the 1980s as a means of stress relief and self-exploration.

### 1.2 Floatation setup

Floatation REST is characterized by an environment designed to minimize external sensory input, thereby facilitating profound relaxation and introspection. The mechanism underlying this technique involves creating a state of sensory isolation, which has been shown to significantly reduce stress and anxiety. Jonsson and Kjellgren demonstrated that participants undergoing Floatation REST experienced notable reductions in stress and anxiety levels, suggesting a direct link between sensory deprivation and mental well-being^17^.

The process of floating in a floatation tank is designed to minimize external sensory inputs, which is achieved through several key features. Visual Minimization: The floatation tank environment is typically devoid of light, reducing visual stimuli. This darkness helps to eliminate distractions from the visual field, allowing the brain to relax^18^. Auditory Reduction: Sound is significantly reduced in a floatation tank. The tanks are often soundproof or designed to minimize external noise. In addition, the participants wear earplugs to further reduce the sound. This lack of auditory input helps to quiet the mind. Tactile Sensations: The water in the tank is saturated with Epsom salt (magnesium sulfate), increasing its density and allowing individuals to float effortlessly. This buoyancy reduces the sense of touch, as there is minimal contact with any solid surface. The water and air are heated to skin temperature, blurring the line between the body and the water ^1^. Thermal Regulation: Maintaining the water and air at skin temperature ensures that the body does not need to actively regulate its temperature, reducing physiological distractions ^19^. Olfactory and Gustatory Inputs: These senses are also minimized in a floatation tank. The environment is kept free from strong smells, and since users are not eating or drinking in the tank, gustatory inputs are absent.

### 1.3 Floatation REST benefits

A significant body of research has focused on the mental health benefits of Floatation REST. For example, Feinstein et al. found a significant reduction in anxiety and improvement in mood among participants, offering a non-pharmacological approach to mental health care ^20^. This finding is supported by earlier studies, which have consistently demonstrated the effectiveness of Floatation REST in reducing symptoms of anxiety and depression ^21^. Floatation REST has also been explored as a treatment for insomnia and stress management. Research suggests that it can improve sleep quality and reduce stress levels, making it a promising option for those with sleep disorders. For example, a study examining its impact on insomnia symptoms highlighted its potential as a treatment for sleep-related issues, with participants reporting improved sleep quality and reduced insomnia symptoms ^22^. The technique has shown promise in managing chronic pain conditions. A study in JAMA Network Open (2021) highlights its effectiveness in reducing pain in chronic pain sufferers, suggesting a potential role for Floatation REST in pain management strategies ^23^. Beyond mental health, Floatation REST has been found to benefit physical well-being, aiding in muscle relaxation and expediting recovery from physical exertion. Therefore floatation REST has gained attention in the field of sports and athletic performance. Athletes have reported enhanced recovery after physical exertion, reduced muscle soreness, and improved mental focus ^24,25^. Neuroscientific research into Floatation REST has begun to shed light on its impact on brain function. The sensory deprivation experienced during floatation can lead to a decrease in cortical activity, allowing the brain to enter a deeply relaxed state similar to that achieved through meditation^26^.

### 1.4 This Study

There are two aims of this study. Firstly, we would like to explore the motivation and people’s perception of the benefits of regular floating sessions practice. Secondly, we would like to gather opinions and suggestions on the focus of the potential future research.

## 2. Methods

### 2.1. Study Designz

This study employed a cross-sectional, descriptive survey to document the following information about individuals who regularly engage in floatation therapy. Centres were (randomly) approached by email where the study was advertised. The participation was based on voluntary consent and was completely anonymous. The following information was requested:

1. Demographics (i.e., age, employment status, and country of residence).
2. Floatation habits (i.e., duration and frequency of floatation sessions).
3. Benefits and side effects (i.e., effects on relaxation, sleep, anxiety, depression).
4. Perspectives on future research directions (i.e., aspects of mental and physical health).

### 2.2. Inclusion/exclusion criteria

Participation was entirely voluntary and anonymous The subsequent questions ensured that only individuals with regular floatation experience participated The initial question enforced the exclusion criterion of being 18 years or older (Answer with ‘yes’ to indicate you have read and understood this information section, and that you consent to participate in this survey and confirm you are 18 years or older.)..

### 2.3. Questionnaire

A descriptive electronic survey comprising 23 questions was developed using Microsoft Forms, with all questions presented in English. The survey included a variety of question formats, such as yes/no, multiple-choice, interval-scale, open-ended, and ratio scale questions. The full questionnaire is available in the Supplementary Materials.

### 2.4. Recruitment

Respondents were recruited via email to floatation centres in various countries, requesting them to inform their clients about our interest in their opinions and experiences. Floatation centres were identified through a search using Google Maps. We contacted 30 centres in the UK, 8 in Ireland, 7 in New Zealand, 17 in Canada, and 40 in Australia. For each contacted Floatation centre we also checked the price for a single float and the best deal available on the webpage.

We cannot state the acceptance rate of participation at the centre level because all data is anonymous. Additionally, we reached out to Michael Cordova, Chair of the UK Float Tank Association and a member of the board of directors of the Float Tank Association. After setting up an online meeting with him, he kindly forwarded our survey details to his contacts. Similarly, we talked to Dr Justin Feinstein, President and Director of the Float Research Collective. Furthermore, we directly approached floatation centres in London, conversed with the staff, and were permitted to display our survey flyer at these locations.

### 2.5. Data Analysis

Microsoft Forms provided an Excel sheet with the survey data. For further statistical analysis, the data from multiple-choice questions required transformation into single 0 or 1 responses for each option. This conversion was executed using the online AI tool, Julius.

We have investigated the whole dataset using clustering – an exploratory, data-driven technique whereby natural, distance-based grouping of participants, may be beneficial for capturing similar traits and characterizing common behaviours of heterogeneous populations or environments. We present the results in terms of silhouette plots-graphical depictions of the individual observations and their corresponding cluster-wide silhouette values for interpretation of the performance and validation of the consistency of the clustering. The silhouette value lies between -1 and 1 and measures the similarity (cohesion) of a data point to its cluster relative to other clusters (separation). For example, a high/low silhouette value indicates that the observation matches its cluster well/poorly. Cluster-wide silhouette values represent the overall averages of the corresponding point-based silhouette values for all members of each class.

We also present the results in terms of bars associated with each variable. The magnitude of the bars represents the coordinates of the cluster centres with higher values of a coordinate indicating the cluster pulled towards that coordinate, i.e. the cluster likely to group people with that particular characteristic.

### 2.6. Ethical Consideration

This study received a favourable ethical opinion from the St George’s Research Ethics Committee. REC Reference Number: 2023.0012.

## 3. Results

In 2004 Dierendonck and Nijenhuis performed a meta-analysis of 27 clinical trials describing the effects of floatation REST on stress reduction as an outcome ^26^. For studies that investigated the difference between pre and post-treatment effects, the overall mean effect size was 1.02, the ones with a shorter treatment time (3 weeks and shorter) showed a smaller effect size of 0.87 and for long-term studies (6 months) the effect size was larger with 1.25. According to Lipsey and Wilson, this shows that floatation REST is in the top 25% of a wide range of anti-stress and coping techniques ^27^. Consequently, many other studies were performed with this technique. However, there is little known about what motivates people to privately pay for the treatment on a regular base. To determine how expensive floating is we checked the websites of all contacted floatation centres (30 in the UK, 8 in Ireland, 7 in New Zealand, 17 in Canada, and 40 in Australia) for the price for a single float or the best deal. The best deals were usually a membership or a package with 3-10 floats. The results of the prices are given in Figure 1.

**Figure 1:**
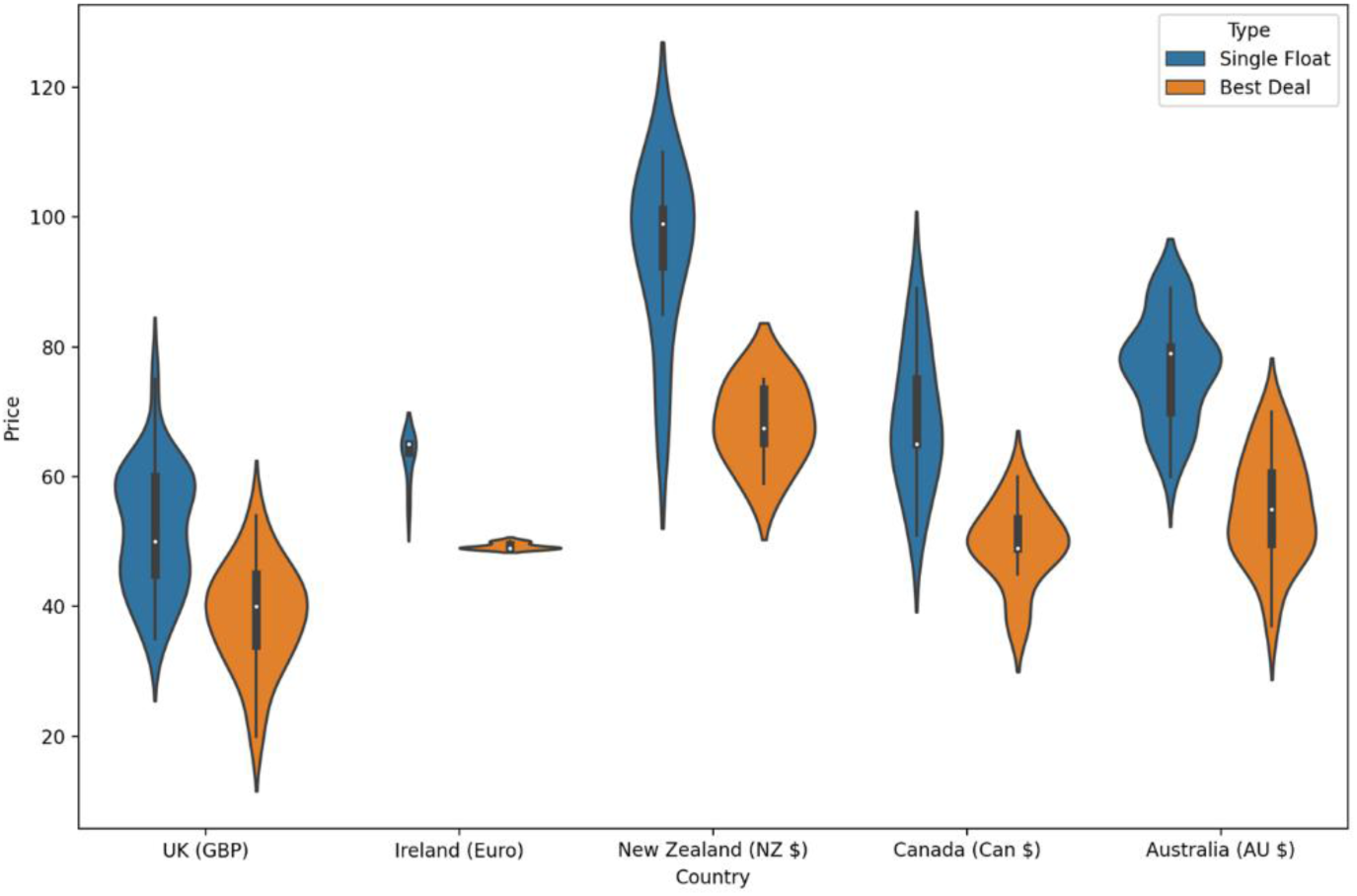
Violin Plot of the prices for a 60-minute float session across all contacted floatation centres, with data collected from the United Kingdom (n=30), Ireland (n=8), New Zealand (n=7), Canada (n=17), and Australia (n=40). The prices were sourced from the websites of these centres in January 2023. Due to variations in currency exchange rates, taxation models, and the cost of living across different countries, prices are presented in the local currency of each country. The primary objective of this analysis is not to compare prices between countries but to highlight the price level, price variations in a country and potential cost benefits of regular floating sessions.

According to the Global Wellness Institute, in 2020, the United Kingdom’s wellness industry was the fifth-largest globally, with a valuation of approximately 158 billion US dollars ^28^. In 2012, the UK spa sector alone attracted over 35 million customers, generating revenue of around 5.2 billion euros from spa treatments and services. When converted using the current exchange rates, this amount averages to about 127 pounds per customer annually. In comparison, a membership for monthly flotation therapy sessions, priced at 40 pounds per session, amounts to an annual expenditure of 480 pounds. This cost is higher than the average yearly spend per customer on spa treatments. However, it is very similar to the average gym membership cost in the UK, which is approximately 44.92 pounds a month ^29^.

Observing that individuals are routinely investing in flotation therapy at a cost comparable to other wellness activities, it’s clear that they perceive a significant value in this service. This pattern of regular financial commitment to flotation therapy underscores a broader interest in understanding the specific benefits that make it appealing. Essentially, the focus is on identifying the unique advantages of flotation therapy that justify its expense in the eyes of its regular users.

In total, there were 83 responses to the survey. The average time required to complete the survey was 13 minutes and 18 seconds. The majority of responses came from the UK (52), followed by Canada (7), Australia (6), New Zealand (5), Ireland (4), Belgium, Serbia and Portugal (each 1) and 1 person did not provide the country information. Further demographic data are presented in Table 1. About two-thirds of participants are female and one-third male, with the majority (70.4%) being between 26 and 55. In addition, about half of the participants are employed and from the 69 people who answered the income question, 60.9% judged their income as medium.

**Table 1.**
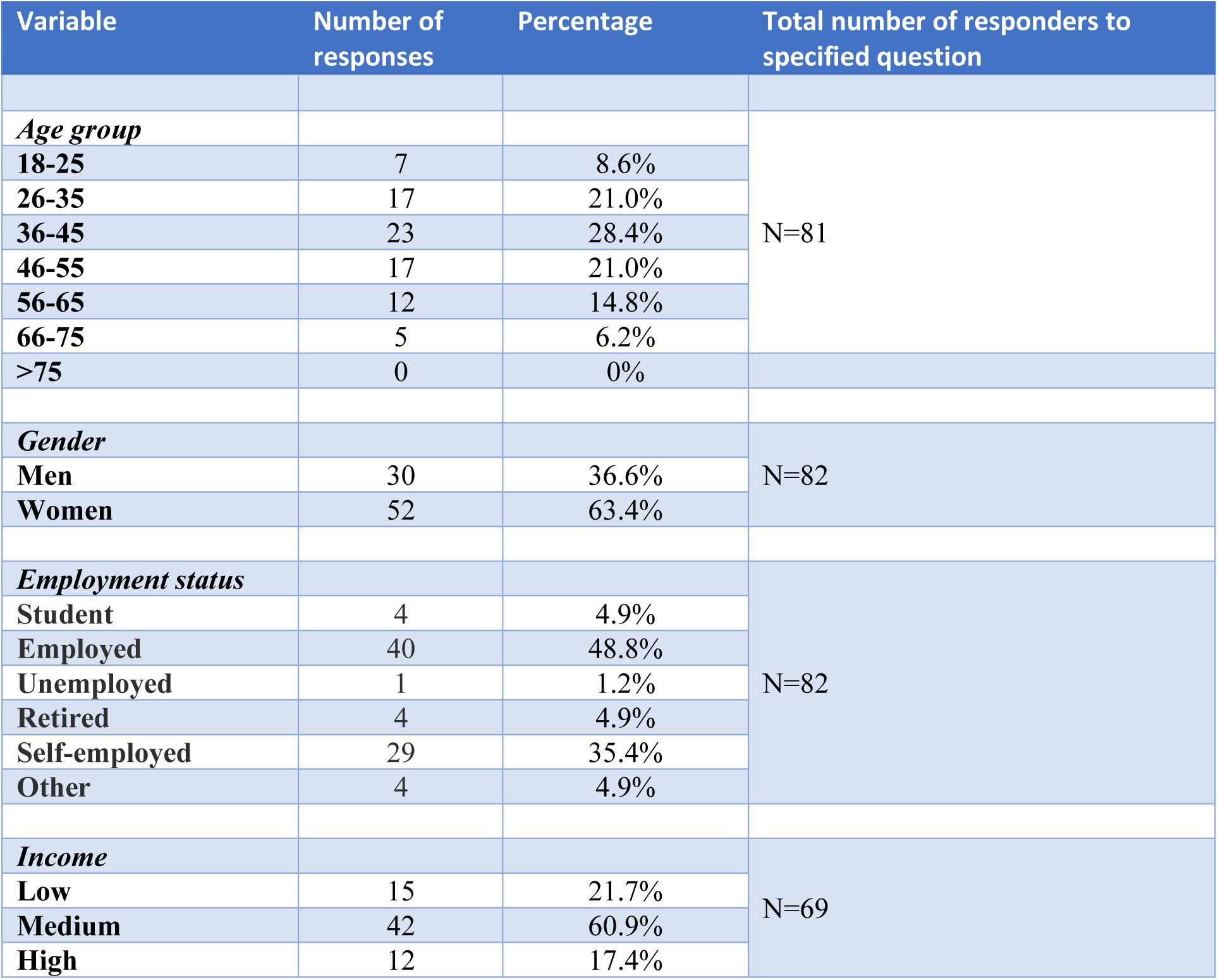
Demographics of participants.

| Variable | Number of responses | Percentage | Total number of responders to specified question |
| --- | --- | --- | --- |
| *Age group* |  |  |  |
| **18-25** | 7 | 8.6% | N=81 |
| **26-35** | 17 | 21.0% |  |
| **36-45** | 23 | 28.4% |  |
| **46-55** | 17 | 21.0% |  |
| **56-65** | 12 | 14.8% |  |
| **66-75** | 5 | 6.2% |  |
| **>75** | 0 | 0% |  |
| *Gender* |  |  |  |
| **Men** | 30 | 36.6% | N=82 |
| **Women** | 52 | 63.4% |  |
| *Employment status* |  |  |  |
| **Student** | 4 | 4.9% | N=82 |
| **Employed** | 40 | 48.8% |  |
| **Unemployed** | 1 | 1.2% |  |
| **Retired** | 4 | 4.9% |  |
| **Self-employed** | 29 | 35.4% |  |
| **Other** | 4 | 4.9% |  |
| *Income* |  |  |  |
| **Low** | 15 | 21.7% | N=69 |
| **Medium** | 42 | 60.9% |  |
| **High** | 12 | 17.4% |  |

More than half of the participants float 1-3 times a month, which coincides with a membership in a floating centre providing better deals for 1 or 2 floats per month. There is a strong desire to float more frequently. If time and money would not be an issue, then more than 50% would float 1-4 times a week and close to 20% even more frequently, see Figure 1.

**Figure 1:**
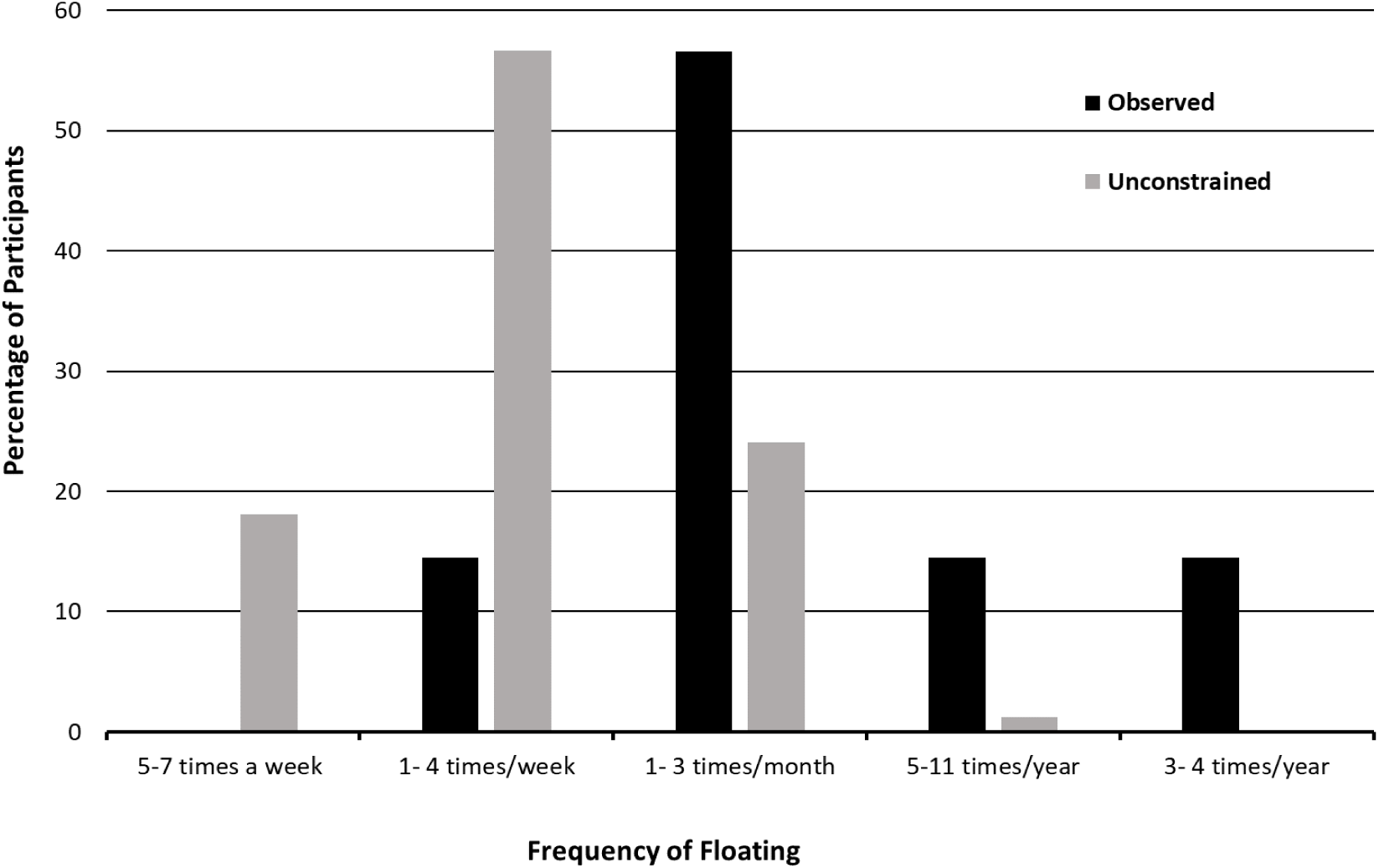
Frequency of Floating. Black shows the frequency reported by the participants. Grey shows the frequency when participants were asked how often they would float if time and money were not perceived as a constraining factor.

In terms of experience with regular floatation, the largest group consisted of individuals with 3-5 years of experience, accounting for 26.5%. Approximately two-thirds of the participants typically engage in 60-minute flotation sessions, aligning with the standard offerings of flotation centres. Notably, 28.9% of participants opt for sessions longer than 60 minutes, with centres commonly providing a 90-minute option. A smaller segment, 6%, exceeds even this duration (refer to Table 2).

**Table 2:** Habits of participants about their floating and satisfaction and support for further floating research.

| Variable | Number of responses | Percentage | Total number of responders to specified question |
| --- | --- | --- | --- |
| *Floating – How long for?* |  |  |  |
| 1 – 4 months | 11 | 13.3% | N=83 |
| 5 – 7 months | 10 | 12.0% |  |
| 1 - 2 years | 15 | 18.1% |  |
| 3 – 5 years | 22 | 26.5% |  |
| 6 – 10 years | 13 | 15.7% |  |
| > 10 years | 12 | 14.5% |  |
| *Duration of typical float* |  |  |  |
| 30 - 60 min | 54 | 65.1% | N=83 |
| 61 – 90 min | 24 | 28.9% |  |
| 91 – 120 min | 3 | 3.6% |  |
| 121 – 180 min | 2 | 2.4% |  |
| > 180 min | 0 | 0.0% |  |
| *Combination with other relaxation techniques* |  |  |  |
| Walks in nature | 39 | 56.5% | N=69, multiple response question |
| Meditation | 37 | 53.6% |  |
| Breath exercises | 36 | 52.1% |  |
| Mindfulness | 29 | 42.0% |  |
| Yoga | 14 | 20.3% |  |
| EFT (emotional freedom technique) | 5 | 7.2% |  |
| Qi Gong | 4 | 5.8% |  |
| Tai Chi Chuan | 3 | 4.3% |  |
| Progressive Muscle Relaxation | 1 | 1.4% |  |
| Others | 17 | 24.6% |  |
| *Satisfaction with floating* |  |  |  |
| 1 | 1 | 1.3% | N=80 |
| 2 | 0 | 0.0% |  |
| 3 | 0 | 0.0% |  |
| 4 | 1 | 1.3% |  |
| 5 | 0 | 0.0% |  |
| 6 | 1 | 1.3% |  |
| 7 | 5 | 6.3% |  |
| 8 | 11 | 13.8% |  |
| 9 | 11 | 13.8% |  |
| 10 | 50 | 62.5% |  |
| *Support for further research on floating* |  |  |  |
| 1 | 1 | 1.2% | N=83 |
| 2 | 1 | 1.2% |  |
| 3 | 0 | 0.0% |  |
| 4 | 0 | 0.0% |  |
| 5 | 0 | 0.0% |  |
| 6 | 1 | 1.2% |  |
| 7 | 2 | 2.4% |  |
| 8 | 5 | 6.0% |  |
| 9 | 6 | 7.2% |  |
| 10 | 67 | 80.7% |  |

Participants were also queried about their practice of combining flotation with other relaxation techniques. The most popular complementary activities included walks in nature (56.5%), meditation (53.6%), breathing exercises (52.1%), mindfulness (42.0%), and yoga (20.3%), as shown in Table 2. Participants reported engaging in various relaxation techniques not explicitly listed in the questionnaire but categorized under “Others.” Notably, two individuals practised sound therapy, three utilized an infrared sauna, one participant used a sauna (type unspecified), and two engaged in light therapy. Additionally, several techniques were mentioned only once, including binaural beats, acupuncture, Bowen therapy, sleep, sea swimming, self-havening, regular massage, and prayer. One person reported no combination with other techniques.

Regarding satisfaction with flotation therapy, participants rated their experience highly, averaging 9.16 on a scale of 1 to 10, where 1 indicates the least satisfaction and 10 the highest. Additionally, there was strong support for further research into flotation therapy, with an average score of 9.48 on a similar 1 to 10 scale, where 1 represents the least support and 10 the most.

The different benefits used in the questionnaire were taken from the literature and the web pages of the floating centres that were contacted. The results of the survey show the prominence of mental health benefits, with relaxation and stress reduction being the most notable, see Figure 2. The vast majority of respondents, 91.6%, reported experiencing relaxation, while 77.1% noted stress reduction. These benefits are integral to mental health, addressing common issues such as stress and tension. This aligns with the holistic understanding that managing stress and achieving relaxation is crucial for emotional and psychological balance.

**Figure 2:**
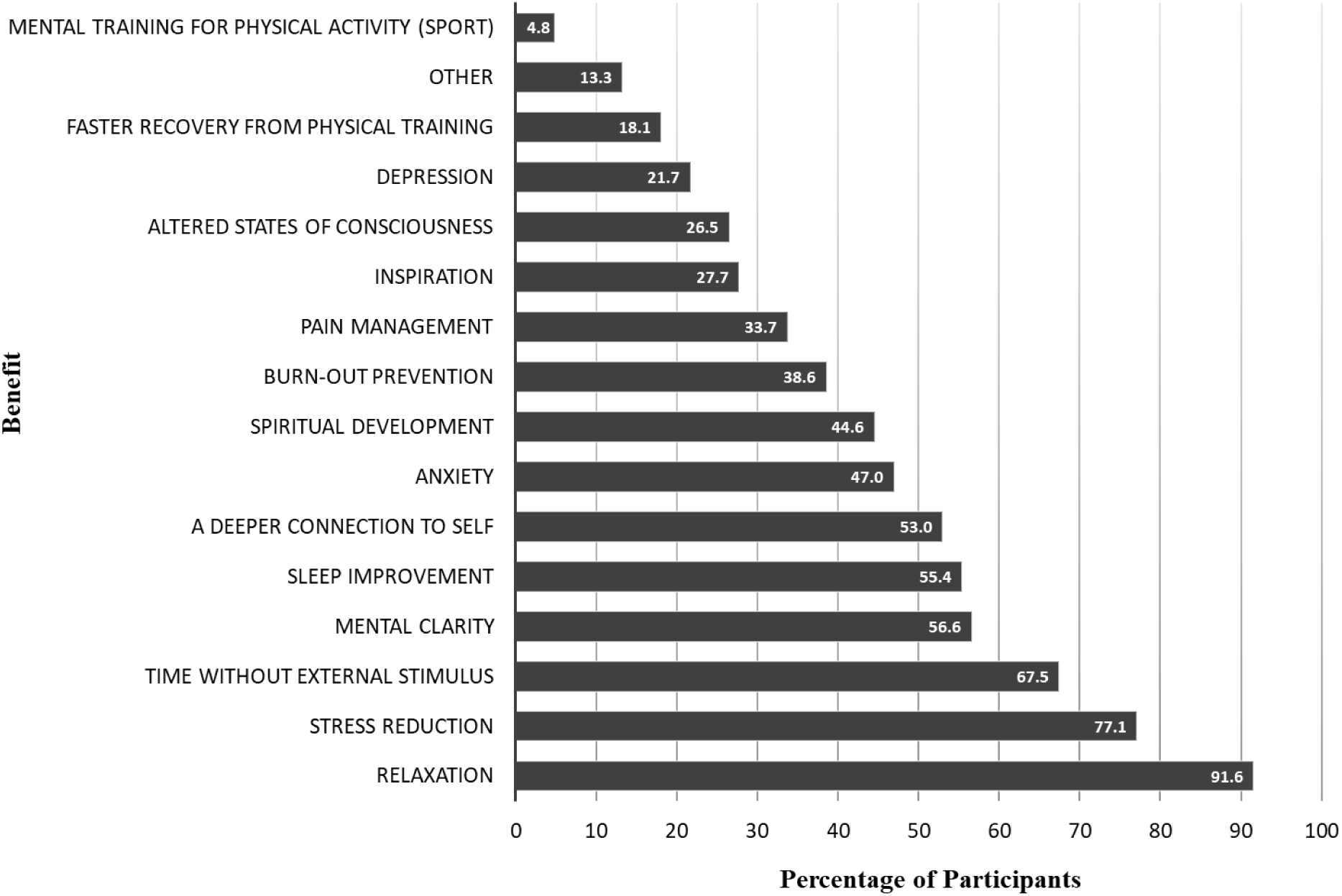
Benefits of floating, multiple answers allowed.

Additionally, 67.5% of participants valued the time without external stimulus, a key aspect of floating that contributes to mental clarity, as reported by 56.6% of respondents. Sleep improvement, a crucial factor in mental and physical health, was noted by 55.4% of participants. The survey also revealed the role of floating in facilitating emotional regulation and introspection, with 53.0% experiencing a deeper connection to self, 47.0% reporting benefits in managing anxiety, as well as burn-out prevention (38.6%) and depression management (21.7%).

Spiritual development was observed by 44.6% of respondents, inspiration was noted by 27.7%, and altered states of consciousness were experienced by 26.5% of respondents, suggesting floating’s capacity to enhance creativity and offer unique psychological experiences.

Less common but still notable were benefits related to physical health and training. Pain management was reported by 33.7% of participants, indicating floating’s role in physical wellness. Faster recovery from physical training was reported by 18.1% of participants, and mental training for physical activity was noted by 4.8%. An additional 13.3% of participants reported benefits categorized under ‘Other’, indicating a range of individualized experiences with floating. Here are some statements captured under ‘Other’: “Mental reboot from demanding job”, “improved energy levels”, “It re-energises me, I can (not always) feel like a new person from the inside out”, “Help with my Complex PTSD”, “Problem solving”, “Its regulated my menstruation cycle”, and “I found this to be one of the most important tool for my well-being instead of yoga or anything like that. I noticed a difference in my mood and energy and emotion without floating”.

An important question was also on how long these benefits last. The majority of participants (53.0%) experienced those benefits for 1 - 4 days, followed by 33.7% where the benefits lasted between 2 - 23 hours. This information can be very helpful for future designs of clinical trials or treatment regimes.

All interventions can have potential side effects. The question about experienced side effects was answered by 55 participants, where 41 (74.5 %) reported no side effects. Three participants reported mind chatter and stress to relax: “mind chatter increases first and therefore stress due to no distractions. Also stress due to the fear it might be over before I gained relaxation”, “Sometimes my mind just refuses to switch off which is very frustrating … does not happen much” and “Not being able to control my mind to fully relax and calm myself”. Another two described issues with their skin: “skin itchiness caused by magnesium deficiency. But not if I float (say) every week.” and “Not really other than really dry skin”. Two other participants described digestive issues: “slight nausea during my very first floating session. Never again after that.” and “I get bad gas”. Also reported was low blood pressure, tiredness and vertigo: “On rare occasions tiredness”, “Only once - my blood pressure dropped after my first float session. I felt very tired - I could’ve slept for the rest of the day - and my pupils dilated.”, “low blood sugar/pressure” and “Vertigo, tight chest”. The remaining three reported pain, spasms or jolts and anger: “Pain as releasing tension in shoulders and head, for a period of time it was every float, now it’s less regular.”, “I once experienced a temporary lack of visual acuity after a float and was unable to focus my sight for about 10-15 minutes, I believe that it was triggered by glancing upwards at bright sunlight coming through a skylight. I also occasionally experience very fast spasms or jolts accompanied by a flash of white light behind closed eyelids while mid float, it’s not unpleasant or causes any discomfort but usually comes right at the edge of losing full consciousness and comes with a mild adrenaline spike” and “Only a couple times, but I have come out stupid angry for seemingly no reason. Feels real lizard brain to me.”.

Participants of this survey were also asked about their opinion about future research concerning floatation therapy, categorized into for aspects: mental health, physical health, body-mind overall health and suggestions, see Table 3. From the three aspects of health, with various categories seven were voted by two-thirds of the participants. In methal health, the majority of respondents (84.1%) recommended research on stress reduction in general. Other significant areas of interest include burn-out prevention, trauma/Post-traumatic stress disorder (PTSD), each receiving 73.2% of responses, and anxiety (70.7%). In physical health pain management was a leading area of interest, with 76.5% of responses. Recovery from injuries and deeper muscle relaxation were also prominent, receiving 74.1% and 65.4% respectively. In body-mind overall health improved sleep was the most recommended area for research, with 82.7% of responses, followed by better self-reflection/self-awareness (67.9%).

Beyond these categories, respondents provided specific suggestions for future research. These included increasing public awareness and dispelling stereotypes about floatation therapy, examining its impact on pregnancy and mental performance, and exploring its effects on chronic health conditions and brain activity during floating. There was interest in investigating the therapy’s role in conjunction with PTSD treatments, in enhancing body confidence, in addiction management, and in facilitating deep relaxation and mind-body-soul connection. Additionally, the potential of floatation therapy in improving outcomes for conditions like skin disorders, migraines, and brain injuries, and in the management of pain without heavy reliance on medication, was underscored. Finally, there was curiosity about the long-term effects of consistent float practice and its comparison with meditation practices and traditional medications.

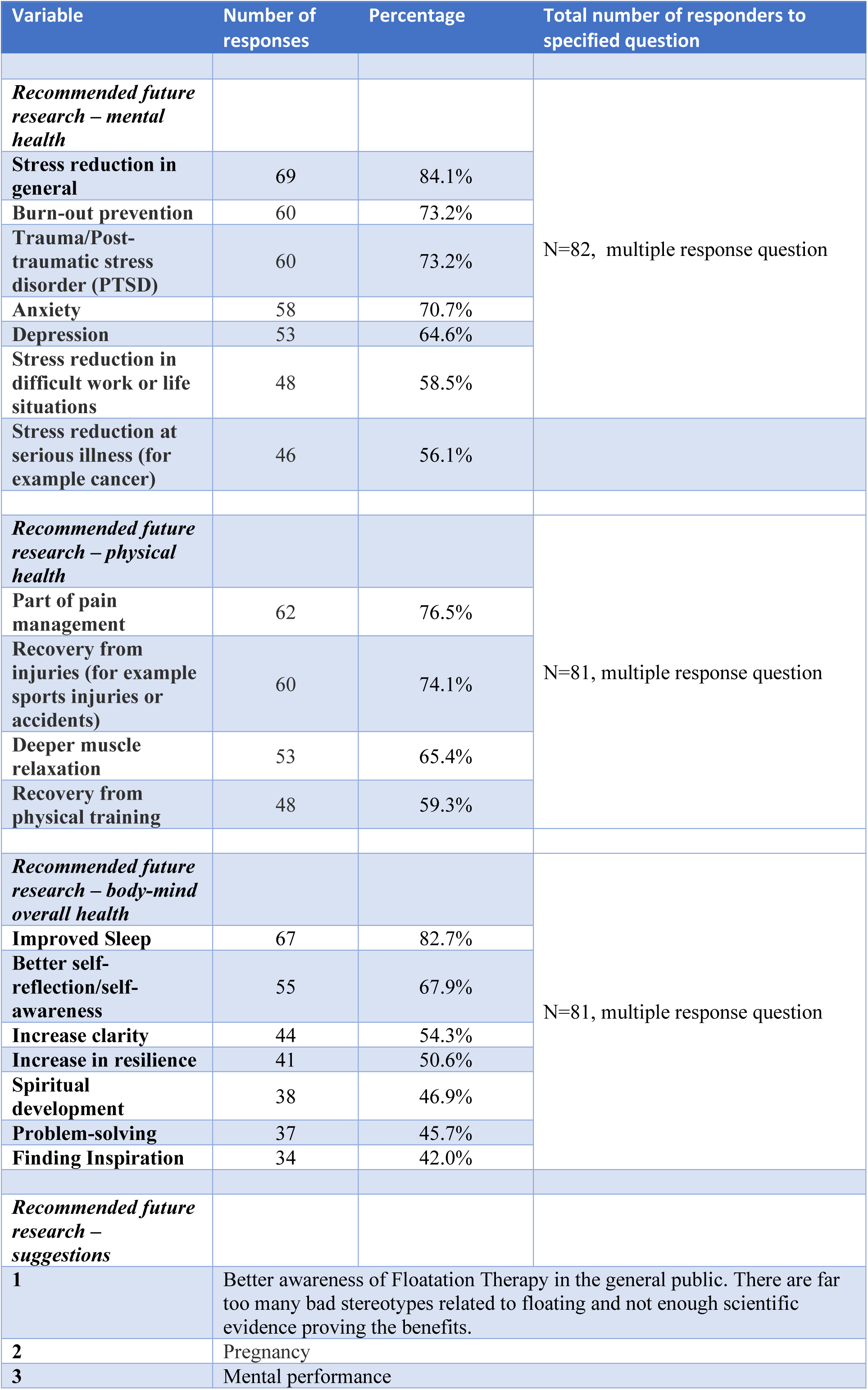
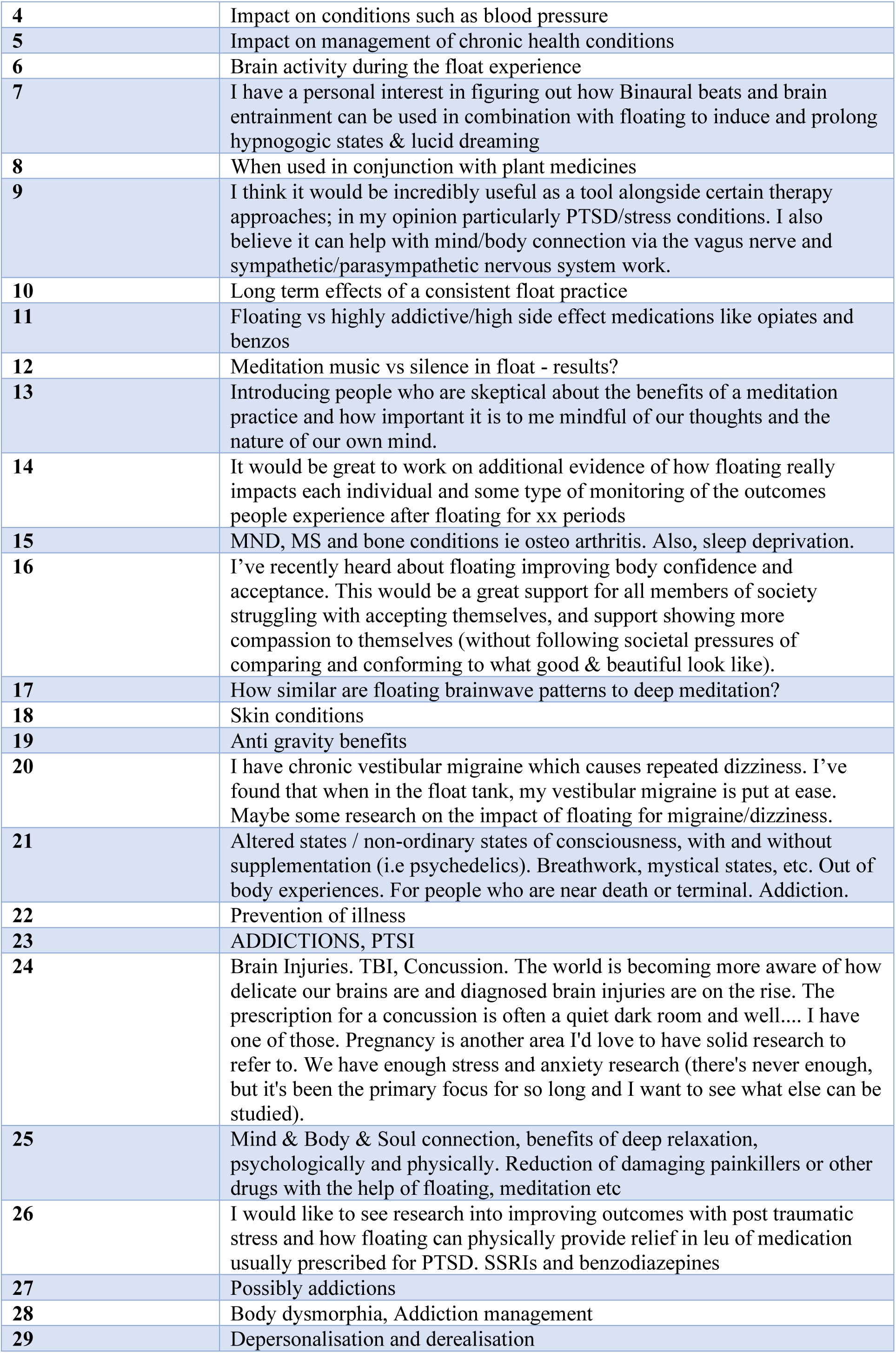

| Variable | Number of responses | Percentage | Total number of responders to specified question |
| --- | --- | --- | --- |
| *Recommended future research – mental health* |  |  |  |
| Stress reduction in general | 69 | 84.1% | N=82, multiple response question |
| Burn-out prevention | 60 | 73.2% |  |
| Trauma/Post-traumatic stress disorder (PTSD) | 60 | 73.2% |  |
| Anxiety | 58 | 70.7% |  |
| Depression | 53 | 64.6% |  |
| Stress reduction in difficult work or life situations | 48 | 58.5% |  |
| Stress reduction at serious illness (for example cancer) | 46 | 56.1% |  |
| *Recommended future research – physical health* |  |  |  |
| Part of pain management | 62 | 76.5% | N=81, multiple response question |
| Recovery from injuries (for example sports injuries or accidents) | 60 | 74.1% |  |
| Deeper muscle relaxation | 53 | 65.4% |  |
| Recovery from physical training | 48 | 59.3% |  |
| *Recommended future research – body-mind overall health* |  |  |  |
| Improved Sleep | 67 | 82.7% | N=81, multiple response question |
| Better self-reflection/self-awareness | 55 | 67.9% |  |
| Increase clarity | 44 | 54.3% |  |
| Increase in resilience | 41 | 50.6% |  |
| Spiritual development | 38 | 46.9% |  |
| Problem-solving | 37 | 45.7% |  |
| Finding Inspiration | 34 | 42.0% |  |
| *Recommended future research – suggestions* |  |  |  |
| 1 | Better awareness of Floatation Therapy in the general public. There are far too many bad stereotypes related to floating and not enough scientific evidence proving the benefits. |  |  |
| 2 | Pregnancy |  |  |
| 3 | Mental performance |  |  |

|  |  |
| --- | --- |
| 4 | Impact on conditions such as blood pressure |
| 5 | Impact on management of chronic health conditions |
| 6 | Brain activity during the float experience |
| 7 | I have a personal interest in figuring out how Binaural beats and brain entrainment can be used in combination with floating to induce and prolong hypnogogic states & lucid dreaming |
| 8 | When used in conjunction with plant medicines |
| 9 | I think it would be incredibly useful as a tool alongside certain therapy approaches; in my opinion particularly PTSD/stress conditions. I also believe it can help with mind/body connection via the vagus nerve and sympathetic/parasympathetic nervous system work. |
| 10 | Long term effects of a consistent float practice |
| 11 | Floating vs highly addictive/high side effect medications like opiates and benzos |
| 12 | Meditation music vs silence in float - results? |
| 13 | Introducing people who are skeptical about the benefits of a meditation practice and how important it is to me mindful of our thoughts and the nature of our own mind. |
| 14 | It would be great to work on additional evidence of how floating really impacts each individual and some type of monitoring of the outcomes people experience after floating for xx periods |
| 15 | MND, MS and bone conditions ie osteo arthritis. Also, sleep deprivation. |
| 16 | I've recently heard about floating improving body confidence and acceptance. This would be a great support for all members of society struggling with accepting themselves, and support showing more compassion to themselves (without following societal pressures of comparing and conforming to what good & beautiful look like). |
| 17 | How similar are floating brainwave patterns to deep meditation? |
| 18 | Skin conditions |
| 19 | Anti gravity benefits |
| 20 | I have chronic vestibular migraine which causes repeated dizziness. I've found that when in the float tank, my vestibular migraine is put at ease. Maybe some research on the impact of floating for migraine/dizziness. |
| 21 | Altered states / non-ordinary states of consciousness, with and without supplementation (i.e psychedelics). Breathwork, mystical states, etc. Out of body experiences. For people who are near death or terminal. Addiction. |
| 22 | Prevention of illness |
| 23 | ADDICTIONS, PTSI |
| 24 | Brain Injuries. TBI, Concussion. The world is becoming more aware of how delicate our brains are and diagnosed brain injuries are on the rise. The prescription for a concussion is often a quiet dark room and well.... I have one of those. Pregnancy is another area I'd love to have solid research to refer to. We have enough stress and anxiety research (there's never enough, but it's been the primary focus for so long and I want to see what else can be studied). |
| 25 | Mind & Body & Soul connection, benefits of deep relaxation, psychologically and physically. Reduction of damaging painkillers or other drugs with the help of floating, meditation etc |
| 26 | I would like to see research into improving outcomes with post traumatic stress and how floating can physically provide relief in leu of medication usually prescribed for PTSD. SSRIs and benzodiazepines |
| 27 | Possibly addictions |
| 28 | Body dysmorphia, Addiction management |
| 29 | Depersonalisation and derealisation |

Cluster analysis serves as a pivotal tool in identifying and interpreting the multifaceted motivations and outcomes associated with flotation therapy among diverse individuals. Cluster analysis, which intricately combines silhouette values and cluster centre components to delineate the diverse motivations and outcomes associated with the therapy. The study identifies three clusters, each with distinct attributes as defined by the analysis, offering a deep dive into the complex landscape of flotation therapy usage.

The silhouette values, which range from -1 to 1, offer insight into how well an individual matches their own cluster in comparison to other clusters, with values closer to 1 indicating a strong match, see Figure 4. For Cluster 1, with 21 members, silhouette values range from -0.015 to 0.18 (average 0.08), suggesting a mix of well-matched individuals and those on the periphery of the cluster’s defining characteristics, see Figure 4. Cluster 2, the smallest group consisting of 9 members, presents silhouette values from -0.04 to 0.12 (average 0.02), pointing towards a relatively cohesive group, albeit with some members having low affinity to the cluster’s core attributes. The largest, Cluster 3, with 51 individuals, showcases silhouette values between 0.04 to 0.29 (average 0.17), indicating the highest degree of cohesion and alignment with the cluster’s defining traits among the three groups.

**Figure 3:**
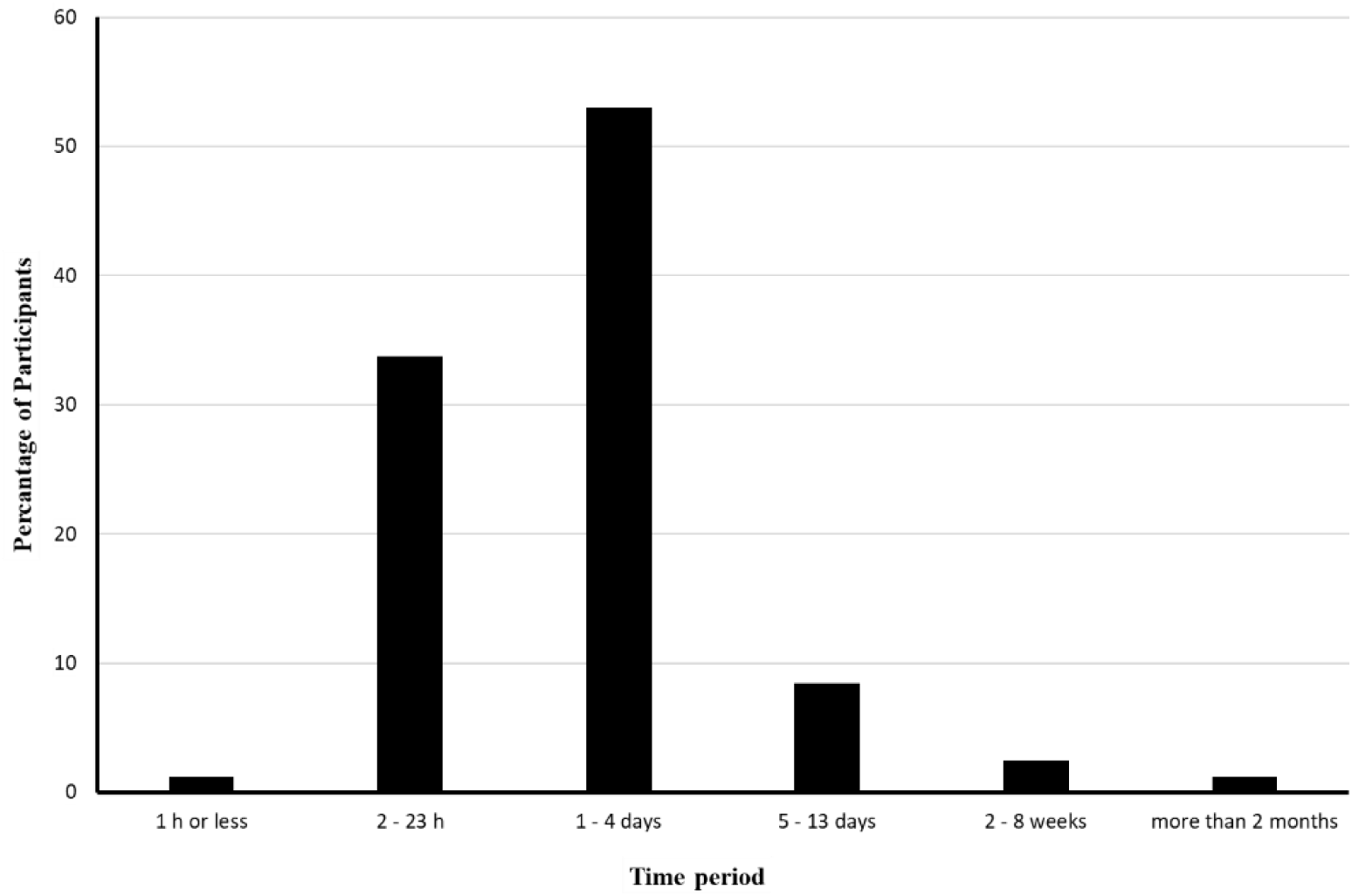
Typical duration of perceived benefits following floatation sessions

**Figure 4:**
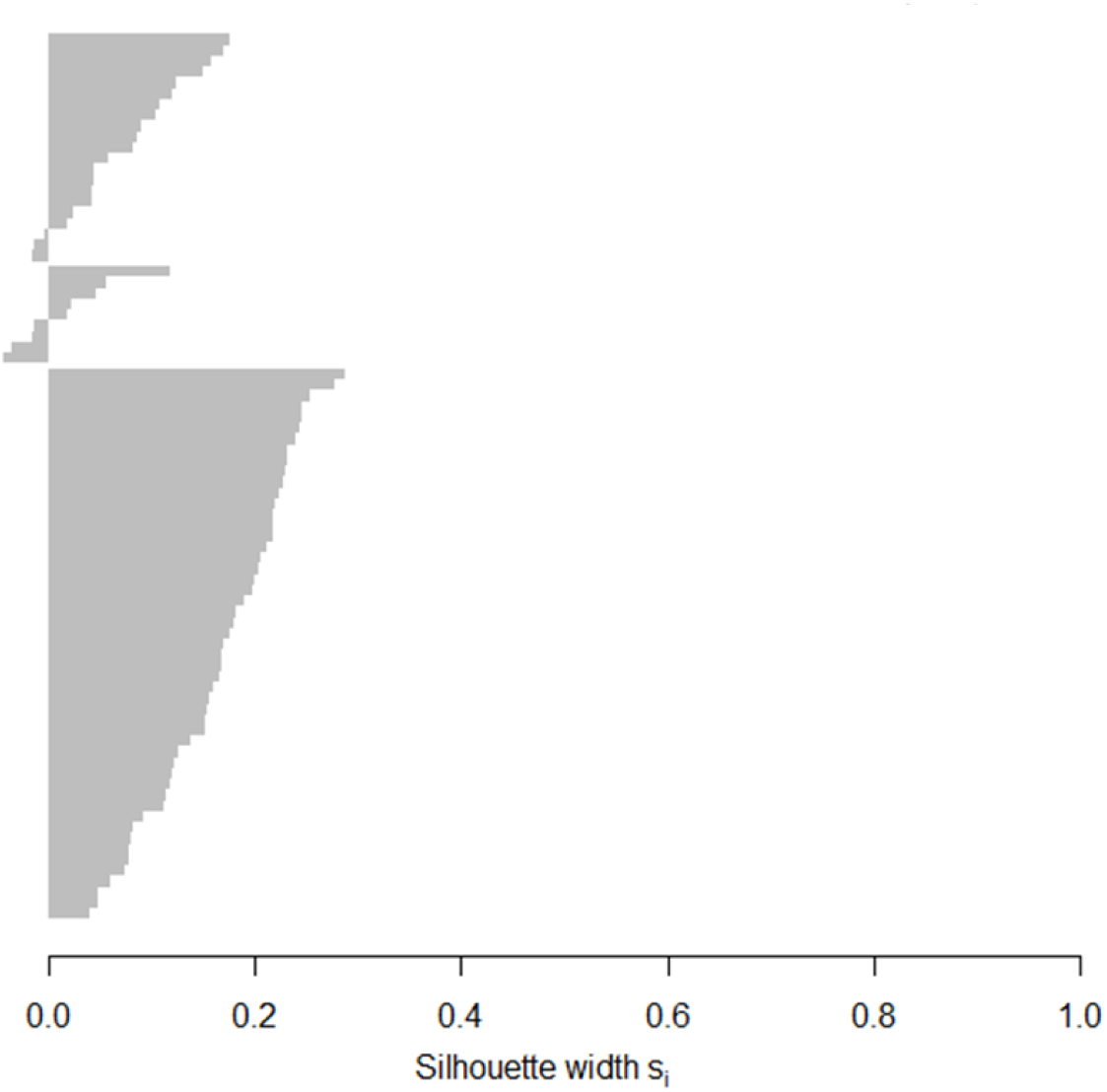
Sillouhette plot indicating how well each observation matches the cluster in which it has been selected. For Cluster 1 (21 members, average of 0.08) the values range from -0.015 to 0.18; for Cluster 2 (9 members, average 0.02) the values range from -0.04 to 0.12 and for Cluster 3 (51 members, average 0.17) the values range from 0.04 to 0.29.

The cluster centre components quantify the average attributes of a cluster’s members, highlighting the prevalent characteristics within each group, see Figure 5. In Cluster 1, the attributes include a high likelihood of seeking inspiration (1.14), a deeper connection with oneself (1.06), experiencing feelings of depression (0.90), engaging in mindfulness practices (0.81), and seeking burnout prevention (0.81). These numbers not only reveal the strength of each characteristic within the cluster but also the intensity with which these traits manifest among its members. Cluster 2 is distinctly marked by a strong engagement in wellness practices such as the Emotional Freedom Technique (1.62), qigong (1.44), taichichuan (1.24), meditation (1.08), and seeking spiritual development (0.90), with high numbers underscoring the significant alignment of its members towards these practices. Conversely, Cluster 3 is characterized by a less likely engagement in meditation (-0.56), seeking spiritual development (-0.51), practising mindfulness (-0.45), seeking connections (-0.46), and engaging in breathing exercises (-0.37), with the negative components indicating these traits are less prevalent among the group’s members.

**Figure 5:**
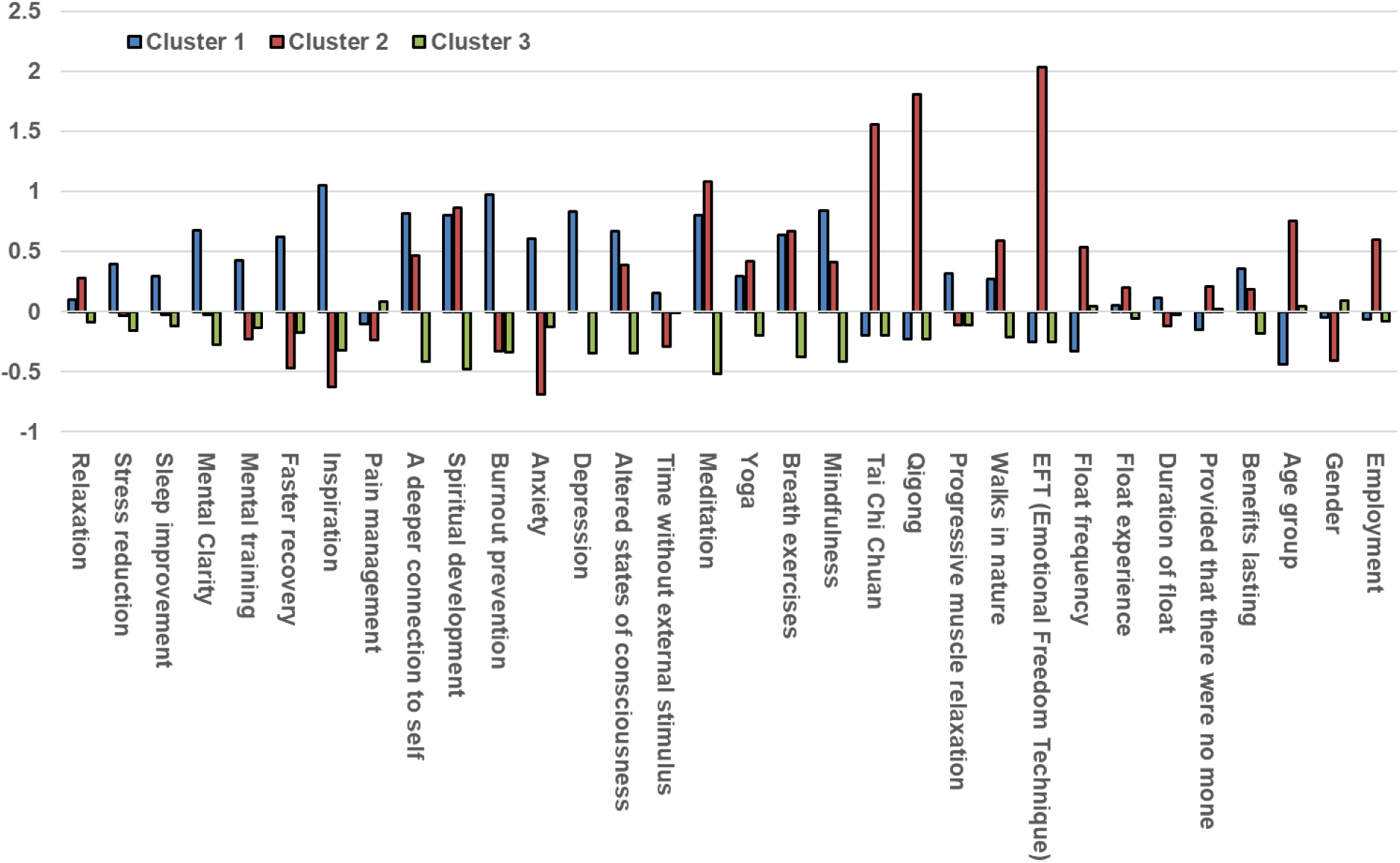
The values of the cluster centers components. A (high) positive value indicate a (highly) likely characteristic; a (large) negative value indicate (highly) unlikely characteristic of the cluster.

This integrated analysis of silhouette values and cluster centre components provides a comprehensive view of how participants are segmented based on their engagement with flotation therapy. Cluster 1 embodies a group seeking mental health benefits and personal development through flotation therapy, despite the variation in silhouette values indicating a range of affinity within the cluster. Cluster 2, although small and slightly cohesive based on silhouette values, is deeply unified by its members’ commitment to specific wellness practices. In contrast, Cluster 3 emerges as the most cohesive group, primarily focused on stress relief and relaxation, as evidenced by both the range of silhouette values and the cluster centre components. Cluster 3’s characteristics suggest a pragmatic approach to flotation therapy, primarily sought for stress relief and relaxation rather than for deeper spiritual or introspective purposes. This detailed understanding highlights the multifaceted appeal of flotation therapy and lays a foundational framework for future research and the development of tailored wellness programs.

## Discussion

The survey conducted in this study was designed with two principal objectives: firstly, to understand why individuals are willing to invest privately in floatation therapy, specifically identifying the benefits they perceive and achieve from this practice. Secondly, the survey aimed to uncover areas where individuals with regular floating experience express a desire for further research.

An extensive outreach was undertaken to gather data from a wide range of floatation centres across various countries. We contacted 30 centres in the UK, 8 in Ireland, 7 in New Zealand, 17 in Canada, and 40 in Australia. Additionally, collaboration was sought with prominent figures in the floatation community, such as Michael Cordova, Chair of the UK Float Tank Association, and Dr. Justin Feinstein, President and Director of the Float Research Collective. Despite these extensive efforts, the study encountered a challenge in its response rate, which introduced limitations in comparing experiences across different English-speaking countries. For instance, in Australia, where 40 centres were contacted, the response rate was unexpectedly low with only 6 responses. This limitation impacts the ability to comprehensively compare and contrast floatation experiences and perceptions among different cultural and geographical contexts. Such a comparison would have been particularly valuable in understanding the diverse reasons behind individuals’ willingness to invest in floatation therapy and if those would differ between different countries. This low response rate highlights the need for future studies to develop more effective strategies for engaging a wider range of participants, especially from regions with a significant presence of floatation centres. This would enable a more robust and diverse understanding of the motivations and desired research directions of floatation therapy users globally.

The survey responses showcased a well-balanced distribution of experience with floatation therapy. This varied range of experience among participants offers a comprehensive perspective on how floatation therapy is perceived and valued over different durations of engagement. The largest group of respondents, consisting of 26.5% with 3-5 years of experience, suggests a significant number of participants have maintained a long-term commitment to floatation therapy, potentially indicating sustained satisfaction and perceived benefits over time. This range of experiences from newcomers to long-term users enriches the study’s findings, offering insights into how the benefits and motivations for floatation therapy might evolve with prolonged engagement.

The participants’ ages varied, with 8.6% aged 18-25, 21.0% between 26-35, 28.4% between 36-45, 21.0% between 46-55, and 20.9% over 55. This distribution indicates that floatation therapy is not confined to a specific age group. While the largest cohort (70.4%) was between 26-55, suggesting an appeal among those likely managing peak career and family responsibilities, the presence of both younger and older adults highlights its broader appeal. The younger adults’ interest might be driven by emerging trends in alternative wellness, whereas older adults might value its potential benefits for pain management, sleep quality, and overall well-being. This diverse age range, along with gender and employment data, underlines floatation therapy’s appeal to a wide spectrum of individuals, influenced by their unique stress experiences and coping mechanisms. The interest in floatation therapy across these demographics highlights its potential as a versatile practice for varied needs and life experiences, reflecting the individualized nature of stress management across different ages ^30,31^.

The study revealed that while more than half of the participants currently float 1-3 times a month, in line with typical floating centre membership plans, there’s a significant discrepancy between this frequency and their desired usage. Over 50% of respondents expressed a wish to float 1-4 times a week, with nearly 20% desiring even more frequent sessions, if not constrained by time and financial factors. This contrast underscores the high perceived value of flotation therapy. However, the cost per session presents a substantial financial barrier, aggravated by the lack of insurance or healthcare coverage despite clinical successes. This situation not only highlights accessibility challenges but also points to broader societal inequalities in accessing wellness therapies. Addressing these disparities is crucial to ensure that the benefits of flotation therapy are available more equitably, calling for systemic changes including healthcare policy reforms and the development of more affordable service models.

Valuable insights into the dynamics of floatation therapy session durations across various income levels were obtained. A detailed breakdown of the participants’ session choices shows that 65.1% opted for float sessions lasting between 30 to 60 minutes, while 28.9% chose sessions between 61 to 90 minutes. Furthermore, a smaller yet notable percentage of participants sought even longer experiences, with 3.6% opting for 91 to 120 minutes and 2.4% for sessions between 121 to 180 minutes. Interestingly, the data challenges certain preconceptions about the influence of financial status on the preference for session length. While it might be assumed that those in the higher income bracket would lean towards longer sessions, perhaps due to a lesser concern over costs and an intention to maximize the value of time invested in commuting to the float centre, the study findings reveal a different trend. A significant majority of high-income participants, constituting 83%, chose the standard 30 to 60 minute sessions. In contrast, of all participants who preferred sessions exceeding 60 minutes, 62% belonged to the medium income group, surprisingly 31% were from the low-income group, and only 7% came from the high-income bracket. There was also a slightly higher preference with men to float longer than 60 minutes (38.7%) compared the women (30.8%). This unexpected pattern of session lengths, especially among the high-income group, implies that personal preferences, perceived therapeutic benefits, and individual lifestyle considerations play a more significant role than financial constraints. The fact that only a small fraction (7%) of the longer sessions were chosen by high-income participants further underscores this point. The study thereby highlights the multifaceted nature of engagement in floatation therapy, where individuals’ choices reflect a complex interplay of economic, personal, and practical factors. This understanding is crucial in the broader context of wellness practices, as it points to the importance of considering a range of motivations and constraints when examining health and wellness behaviours. It also emphasizes the need for more inclusive and diverse approaches in wellness services to cater to varying needs and preferences across different demographic groups. This nuanced view challenges simplistic assumptions about the role of financial capacity in determining health and wellness choices and underlines the necessity for a more comprehensive understanding of individual engagement in therapies like floatation.

The integration of flotation therapy with other relaxation techniques, most of which are free of charge, highlights a comprehensive approach to wellness. Popular practices such as walks in nature (56.5%), meditation (53.6%), breathing exercises (52.1%), mindfulness (42.0%), and yoga (20.3%) complement flotation, possibly enhancing its benefits. This tendency to combine paid therapy like flotation with cost-free activities suggests a balanced approach to well-being, where participants leverage both specialized treatments and accessible practices to achieve holistic health benefits.

One of the main results was identifying the benefits of the participants regarding their floating sessions. The study vividly illustrates flotation therapy’s crucial role in addressing the mental health challenges of our fast-paced, high-stress world. The high prevalence of participants reporting benefits such as relaxation (91.6%) and stress reduction (77.1%) highlights the therapy’s effectiveness in providing a much-needed mental respite. In an age where digital overload and constant connectivity are the norms, 67.5% of participants valued the sensory isolation offered by flotation therapy. This indicates a growing demand for environments that allow disconnection from external stimuli, emphasizing the importance of solitude and quiet in mental well-being. The benefits of flotation therapy extend beyond mere relaxation, with 56.6% of participants experiencing enhanced mental clarity and 55.4% reporting improved sleep quality. These figures reflect the therapy’s broader impact on cognitive and restorative functions, which are often compromised in today’s hectic lifestyle. Interestingly, while the physical health benefits, such as pain management (33.7%) and aiding recovery from physical training (18.1%), are recognized, they were less emphasized among this group of participants. This could suggest that the primary appeal of flotation therapy for these individuals lies in its mental health benefits, rather than its physical health applications. This could also suggest a possible underutilization or under-awareness of the physical benefits of flotation therapy among participants, despite existing research highlighting its efficacy in both mental and physical domains ^32^. The contrast between the study’s findings and the broader research landscape points to the need for further exploration into why participants may prioritize mental health benefits and how awareness of flotation therapy’s full range of benefits can be increased.

Another notable finding was that 53.0% of participants reported benefits lasting 1-4 days and 33.7% for 2-23 hours. That is in agreement with a study Garland et al., demonstrating benefits of flaotation rest for 48 hours for depression and anxiety^33^. That coupled with the expressed desire for more frequent sessions, opens up a significant area for further exploration in flotation therapy research. This desire for increased frequency, alongside the sustained benefits reported, raises important questions about the cumulative effects of flotation therapy. In disciplines like meditation, the depth and extent of benefits are known to intensify with more frequent practice. In flotation therapy, it remains unclear whether more frequent sessions would not only prolong but also intensify the benefits experienced. Future clinical studies, therefore, should consider experimenting with more frequent sessions, possibly twice a week or more, to delve deeper into these potential cumulative effects. This approach could unveil deeper insights into the lasting impact of flotation therapy, its efficacy in enhancing mental and physical well-being, and the optimal frequency required to achieve maximum sustained benefits.

The participants’ strong support for further research into flotation therapy is obvious. Their key areas of interest for future studies include stress reduction (84.1%), burn-out prevention, trauma/PTSD, and anxiety, each receiving substantial support. Pain management (76.5%), recovery from injuries, and deeper muscle relaxation are also prominent areas of interest. Furthermore, improved sleep (82.7%) and better self-reflection/self-awareness rank high on their research agenda. Suggestions to increase public awareness about flotation therapy, its impact on pregnancy and mental performance, and its effects on chronic health conditions and brain activity highlight the engaged and participatory nature of this community. This community-driven approach to research is vital, ensuring that future studies are aligned with the real-world experiences and needs of those who use and benefit from flotation therapy.

The cluster analysis conducted within the “Submerging into Serenity” study provides a distinct categorization of regular flotation therapy users, revealing their motivations and the perceived benefits they associate with the practice. This analysis utilized silhouette values to gauge how well individuals align with their assigned clusters. Cluster 1, with silhouette values ranging from -0.015 to 0.18, signifies a group of 21 individuals with moderate cohesion, who engage in flotation therapy for personal development and mental health benefits, as indicated by their affinity for inspiration (1.14), self-connection (1.06), and mindfulness (0.81). Despite some variance in silhouette values, which might suggest varying levels of alignment within the cluster, there is a marked leaning towards using flotation therapy as a holistic wellness tool.

Cluster 2, the smallest with 9 individuals, has silhouette values between -0.04 to 0.12. It is characterized by members who are highly engaged in wellness practices, such as the Emotional Freedom Technique (1.62), qigong (1.44), and meditation (1.08), revealing a dedicated approach to combining flotation therapy with a broader wellness regimen. Although the silhouette values suggest only slight cohesion, the high numbers for specific practices imply a significant shared interest among its members.

The largest, Cluster 3, consists of 51 members and has silhouette values from 0.04 to 0.29, indicating the strongest cohesion and uniformity in their approach to flotation therapy. This group is distinguished by its less likely engagement with activities typically associated with introspection or spirituality, such as meditation (-0.56) and mindfulness (-0.45), highlighting a pragmatic orientation towards flotation therapy primarily for relaxation and stress relief rather than for deep self-exploration or spiritual development.

These insights provide a rich tapestry of how different segments of the population utilize flotation therapy to cater to their unique wellness needs. The segmentation revealed by the cluster analysis not only underscores the multifaceted appeal of flotation therapy but also furnishes a detailed understanding that enriches the manuscript’s contributions to the discourse on flotation therapy’s role in wellness and personal development. It offers a foundation for future research directions and the development of customized wellness interventions, aiming to enhance the efficacy and appeal of flotation therapy for various user profiles.

To what other method can floating therapy be compared? Maybe the most similar technique is meditation. There are various meditation methods but most of them also lead to less awareness of the environment of the practitioners but rather to the inside. In addition, the stream of thoughts will be reduced in most practises, therefore creating a restricted sensory environment by changing awareness. To no surprise the benefits are similar, they include positive changes for anxiety, depression, sleep distortion, high blood pressure and anti-inflammatory effects ^34^. However, the benefits of meditation are seen with long-term practice and being restricted to a defined position over a longer period of time can lead to tension in the body^34^. Floating on the other hand leads to a very strong relaxation of the muscles often followed by relaxation of the mind by creating a restricted sensory environment. It is unclear how long term exposure to this treatment will change the benefits.

The conclusion of this study is the significant potential of flotation therapy as a wellness tool, particularly for mental health benefits like stress reduction and relaxation. The study reveals a strong desire among participants for more frequent sessions, suggesting that the full benefits of flotation therapy might be better harnessed with increased frequency. Participants’ recommendations for future research indicate a community-driven approach, focusing on areas such as pain management, sleep improvement, and mental health conditions like PTSD. These conclusions underscore the need for more comprehensive research and public awareness to fully realize and utilize the therapeutic potential of flotation therapy.

## Data Availability

All data produced in the present study are available upon reasonable request to the authors

## Sublemental Material

Survey Questions

- For how long are you floating?
  ○ 10 years; >5 years, >2 years; >1 year, > 6 months < 6months
- How regular are you floating on average?
  ○ 2-3 per week, > 2-3 per week, 1 per week, 1 every second week, 1-2 per month; 1 every second month, 1-4 times a year
- Provided that there were no money or time restrictions, how many times would you like to float?
  ○ 1 per day; 1 per day, 6-4 times a week, 3-2 per week, 1 per week, 1 every second week, 1-2 per month; 1 every second month, 1-4 times a year
- What are the main benefits from floating that makes you coming back? Tick all boxes that apply. Important: please fill in others if you have benefits outside these examples to inform us about those.
  ○ Relaxation; improvement in sleep; mental training; faster recovery from physical training, stress reduction, inspiration, problem solving, pain management, illness, mental clarity, deeper connection to self or, deeper connection to others, spiritual development, burn-out prevention, anxiety, depression, altered states of consciousness, time without external stimulus, others
- In you experience or opinion, how many times a week/month is optimal to experience benefit?
  ○ 1 per day; 1 per day, 6-4 times a week, 3-2 per week, 1 per week, 1 every second week, 1-2 per month; 1 every second month, 1-4 times a year
- How long last the benefits after the floating session?
  ○ 1 hours or less, a few hours, 1 day, 2-4 days, 5-7days, 1 week, 2 weeks, 3 weeks, 4 weeks, 2 months, >2 months
- Are you combining floating with other techniques (for example meditation, breathing exercise, mindfulness)?
  ○ Please state this here
- Considering floating and further research, on what topic would you recommend future research should focus on? We provided some topics, but please fill in the others box to inform us better.
  ○ Mental health (for example anxiety, depression), Recovery from injuries (for example sport injuries or accidents), Part of pain management, Recovery from physical training, Mental training, Stress reduction in general, Stress reduction in difficult work-life situations, Stress reduction for serious illness (for example cancer), Burn-out prevention, Deeper relaxation; Better sleep; Finding Inspiration; Problem solving, Increase mental clarity, Increase in resilience; Better self-reflection/self-awareness; Spiritual development, others
- What country are you living in?
- What age group are you belonging to?
- Are you male, female, transgender ….?
- Would you consider yourself earning in low, middle or high income class?

